# Time trends and area-level socioeconomic inequalities in early childhood development at 2 to 2.5 years old in England between 2019 and 2024

**DOI:** 10.1101/2025.03.14.25323968

**Authors:** Yu Wei Chua, Caitlin Murray, Luke Munford, Davara Bennett, Dougal Hargreaves, David Taylor-Robinson

## Abstract

**Importance:** Internationally, there are gaps in data to monitor both early childhood development (ECD) and progress in closing the inequality gap. The unequal impact of the COVID-19 pandemic, and any differential impact on ECD domains is also not poorly understood.

**Objective:** To examine time trends and area-level socioeconomic inequalities in ECD at 2 2.5 years in England between 2019 and 2024

**Design:** Cross-sectional and longitudinal ecological analysis

**Setting:** 149 local authorities in England

**Participants:** 143 local authorities (662 annual observations, publishable quality data and at least 75% coverage of eligible children)

**Exposure:** Year of assessment, area-level socioeconomic conditions (Index of Multiple Deprivation 2019, Income Deprivation Affecting Children Index (IDACI)) assessed as the Slope Index of Inequality (SII) and Relative Index of Inequality (RII))

**Main outcomes:** Ages and Stages Questionnaire 3, rate per 100 not developmentally on track (Five domains of development: Communication, Gross Motor, Fine Motor, Personal Social, or Problem Solving; and any domain)

**Results:** Rate per 100 children not developmentally on track in any domain increased progressively from 2019 (16[95%CI: 14.2; 16.9]), and was highest in 2023 (20.6[18.9; 22.4]) and 2024 (19.8[18.2; 21.4]). Compared to 2019 rates, the largest absolute increase was in 2023 (4.9[3.6; 6.3]), largest for Communication (4.3[3.5; 5.2], followed by similar increases for Personal Social (3.3[2.5; 4]) and Problem Solving (2.9[2.2; 3.5]), and smaller increases for Fine Motor (1.4[0.6; 2]) and Gross Motor (0.9[0.2; 1.6]). All rates except Gross Motor remained elevated in 2024. On average, 21.3[19.5; 23.0] per 100 children were not on track in any domain for the most income-deprived quintile compared to 16.2[14.7;17.8] for the least income-deprived (SII: 2.9[1.8; 3.9]; RII: 23%[14%; 32%]). Inequalities were largest in Communication (SII: 2.8[2.0; 3.6]; RII: 36%[25%; 49%]). Year by area-level socioeconomic conditions interaction effects were not statistically significant.

**Conclusions and Relevance:** In England, ECD worsened during the pandemic, more so for children exposed for longer, or from a younger age. Children born after the pandemic continue to be affected. Area-level inequalities were striking but did not worsen during this period. Pandemic-recovery efforts need to consider the potentially enormous economic and societal cost of disruption to ECD.

**Key points:** *Question:* What are the time trends and area-level socioeconomic inequalities in early childhood development at 2 to 2.5 years in England around the time of the COVID-19 pandemic?

*Findings:* Early childhood development at 2 to 2.5 years in England worsened in the wake of the COVID-19 pandemic. Stark socioeconomic inequalities were observed throughout the period of 2019 to 2024.

*Meaning:* Policy makers need to prioritise early years and children services in pandemic recovery efforts to improve early developmental outcomes, especially for children from socioeconomically deprived backgrounds.

## Introduction

Every child has a right to develop to their full potential, but socioeconomic inequalities in health and development have been evidenced from conception to adulthood.^1^ Globally, poverty, stunting and malnutrition disrupt children’s development.^2^ In high income countries, rising child poverty and its impact on children’s development is a growing concern.^3^ In the UK, inequalities in poverty, school readiness, and other health and educational outcomes across childhood were widening before the pandemic. Promoting early childhood development (ECD) is a foundation for all children to reach their full potential and a priority for sustainable development.^2,4–7^ In particular, the first 1000 days (period of conception to two to three years of life), can be seen as a window of opportunity to improve lifelong outcomes due to the rapid rate of neural development during this time.^2,6,8^

There are gaps in national and cross-national data and measurement tools to track ECD.^9^ Monitoring progress in ECD outcomes at a population-level helps support financial and political commitments to improve ECD, and monitor the impact of policies, programmes and investment in early childhood.^10^ Despite extensive evidence from birth cohort studies showing the detrimental impact of socioeconomic deprivation on ECD,^9^ up-to-date, population-level data is needed to track progress in closing inequalities, and provide intelligence for policymakers on the children most in need of early support.

To inform recovery efforts after the COVID-19 pandemic, it is imperative that we understand the extent to which the pandemic impacted children’s development, including any worsening inequalities in ECD. Experts had warned during the COVID-19 pandemic that household economic shocks, impact on caregiver stress and ability to provide nurturing care, and disruption of services, can disrupt ECD.^11^ Evidence now shows that preschool children and infants born during, or exposed to the COVID-19 pandemic were delayed in their development, compared to children not exposed to the pandemic.^12–14^ However, some studies have suggested that certain domains may be unaffected, and that communication development may be most affected.^15–17^ Evidence of whether preexisting socioeconomic inequalities in ECD were exacerbated by the pandemic is limited and mixed.^18–21^

In the UK, population-level indicators of ECD have previously relied on indicators of school readiness at school entry. Recently available data on ECD assessed at 2 to 2.5 years old now enables monitoring of outcomes nationally, just after the critical first 1000 days. Child-level data is submitted by health visitors to the Community Services Dataset, but most of the data is not yet research ready.^22^ No study has examined the area-level aggregated data available in the interim.^23^ Using area-level estimates, this study assesses national time trends and socioeconomic inequalities in the rates of children not developmentally on track at 2 to 2.5 years in five domains of development, and in any domain, in England around the time of the COVID-19 pandemic.

## Methods

We report an ecological area-level analysis assessing longitudinal trends in children not developmentally on track at 2 to 2.5 years between 2018/19 to 2023/24 (hereafter 2019 to 2024) in England, the extent of socioeconomic inequalities and whether inequalities changed during this period.

### Data sources and measures

Outcome data from 149 local authorities (LA), after accounting for LA boundaries and boundary changes, were obtained from the Health Visiting Service Delivery Metrics (HVSDM), published by the Office of Health Improvement and Disparities (Appendix S1).^24^

ECD at 2 to 2.5 years was measured using the Ages and Stages Questionnaire 3 (ASQ-3),^25^ as part of universal mandatory developmental reviews in England carried out by Health Visitors.^26^ The ASQ-3 screens for children requiring further developmental evaluation, based on cut-off scores of expected development in five domains: Communication, Gross Motor, Fine Motor, Problem Solving and Personal Social Skills. We obtained to the rate of children *not* developmentally on track, per 100 children assessed on the ASQ. Data on the number of children developmentally on track submitted to the HVSDM has been validated to locally held data.^20^ Data from seven assessment years (2018 to 2024) were available at the time of analysis.

The percentage of children receiving developmental reviews was over 90% in all years, but lower for children receiving an ASQ-3 review.^27^ As the ECD metric was classed as experimental statistics up to 2021 due to data quality and coverage of the ASQ review,^24^ we obtained data on the quality (number of LAs submitting publishable quality data) and coverage of ASQ-3 review (percentage of eligible children receiving an ASQ-3 review).

Our exposures were assessment year and childhood socioeconomic conditions (SECs). SECs was measured using the Income Deprivation Affecting Children Index (IDACI) Average Score from the 2019 Indices of Multiple Deprivation.^28^ This score measures the proportion of children experiencing income deprivation in the LA, averaged across deprived and non-deprived areas. A higher score indicates greater income deprivation. We ranked LAs and created quintiles where a fifth of the 2019 child population fell into each income deprivation quintile. A higher quintile indicates lower income deprivation. We converted the ranks into a continuous score ranging from 0 (most advantaged) to 1 (least advantaged) to investigate the Slope and Relative Index of Inequality (SII and RII), such that model parameters capture the absolute and relative difference in children not developmentally on track in the least compared to the most advantaged.

We identified covariates using Directed Acyclic Graphs (DAGs) to isolate the relationship of interest and address potential selection bias (Appendix S2). Children of non-White ethnicity were previously found to be less likely and children from the most deprived decile more likely to receive an ASQ review, in 2019 and 2020 data.^20^ Selection bias can occur if these factors, and any unobserved factors affecting review, are also associated with the outcome (Appendix S3). The adverse effects of COVID-19 lockdown may be biased by the background trend of generally improving employment rates in households with children.^29^ As our focus was on time trends related to COVID-19 impact, we controlled for ethnic composition (Percentage of 0 to 15-year-old children of White British/Irish ethnicity from the 2021 UK Census)^30^, income deprivation (IDACI), ASQ coverage to proxy for other unobserved factors affecting review, and employment rates at birth (Annual Population Survey/Labour Force Survey, households with children 0 to 15 years, by combined economic activity status^31^). In analyses of childhood SECs, we controlled for Year, ASQ coverage and ethnicity.

### Statistical analyses

Analyses were carried out in R (V4.3.2). We preliminarily described data quality and coverage over time to inform main analyses (Appendix S3, Figure S1-S2). Due to high positive skewness and kurtosis of the outcome (Table S1), we trimmed the dataset, log-transformed the outcome and checked regression assumptions (Appendix S1, Figures S3-S7).

In main analyses, we first obtained means and 95% confidence intervals of each outcome over time, and visualised time trends by income deprivation quintile.

Second, using linear mixed-effect models (Appendix S3), we estimated associations of Year and SECs on rates of children not on track in (1) any domain and (2) in each of the five domains. Analyses of Year controlled for ASQ coverage, ethnic composition, IDACI, and employment rates at birth as fixed effects. We entered Year as a categorical variable, to model a complex exposure capturing annual trends, and exposure to COVID-19 shocks (e.g. lockdowns, impact on the economy). Analyses of SECs used continuous IDACI rank as the exposure, controlling for Year, ASQ coverage and ethnic composition in linear mixed-effect models. Interaction effects of SECs by Year were examined using Likelihood Ratio tests. All Models included random effects term to represent normally distributed means of each LA.

Third, we exponentiated model coefficients for the RII. For the SII, we obtained the difference in predicted values between the least advantaged versus most advantaged, for each year. 95% confidence intervals were generated using 1000 nonparametric bootstrap samples of the original data.

### Service-use and economic costs

Referring to population estimates and cost estimates in the published literature, we estimate potential contemporaneous and long-term implications of the increase in children not on track in ECD, on service use and economic losses (Appendix S1 and Table S2). We focus on key domains of speech and language therapy, special educational needs, parental employment losses, and projected implications on workforce participation and labour productivity.^32–34^

## Results

### Sample characteristics

Availability of publishable quality data was poor in 2018, relative to latter years, which had comparable levels of data: 74 (49.0%) LAs submitted publishable ASQ data in 2018, 113 (74.8%) in 2019 and rising to 138 (91.4%) in 2024 (Table S3). IDACI average score was correlated with ASQ coverage only in 2019 (*τ*=-0.12, p=0.032), indicating that coverage was better in areas with lower levels of income deprivation, but not in other years (Table S4). Most LAs achieved over 90% ASQ coverage, but there was variability in coverage with very low coverage in some LAs or year (Table S5). We proceeded with analysing data from 2019 onwards, with at least 75% ASQ coverage (n=143 (96.0%) LAs) (Table S6 and Figure S8).

### Time trends in early childhood development

Rates per 100 children not on track in any domain increased over time (Figure 1), from 15.6[95%CI: 14.2; 16.9] in 2019, peaking in 2023 at 20.6[18.9; 22.4] and decreasing slightly to 19.8[18.2; 21.4] in 2024 (Table S7). Rates were highest for Communication across time.

**Figure 1.**
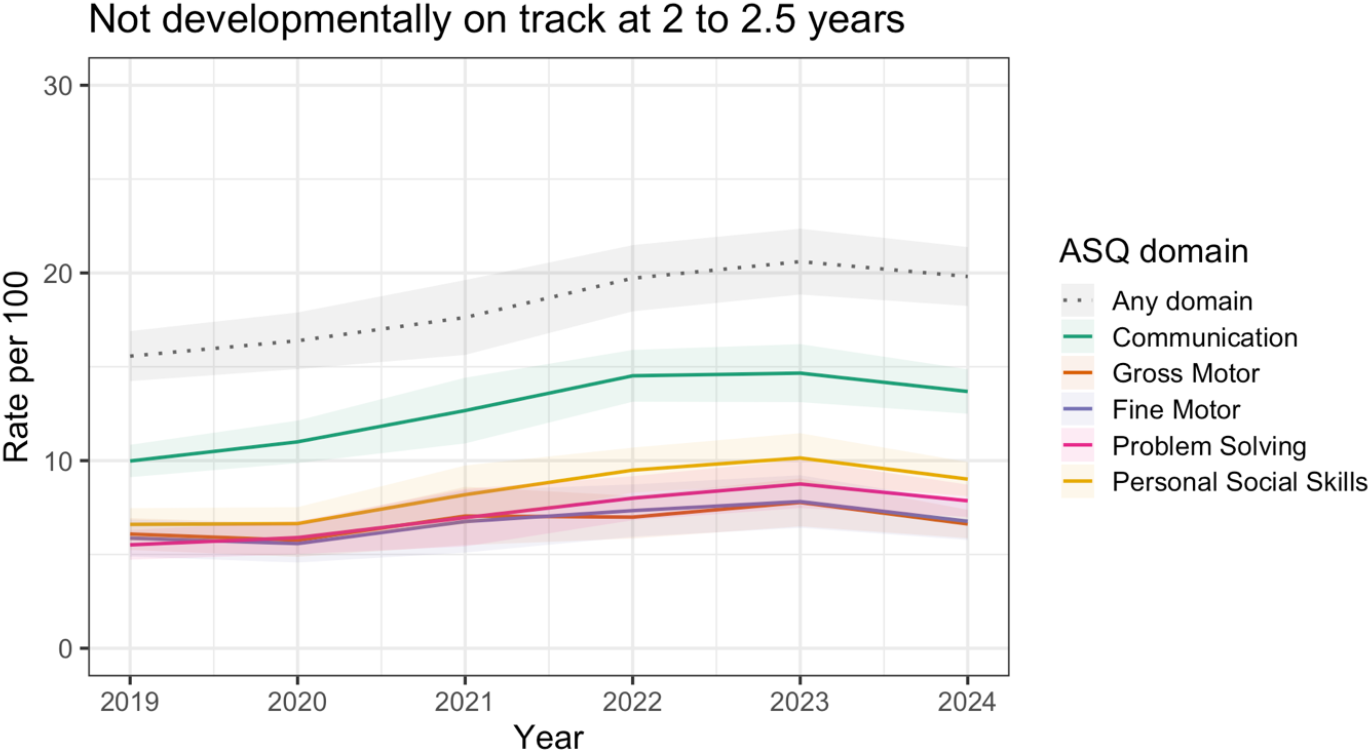
Mean and confidence intervals of children not developmentally on track in any domain and each domain over 2019 to 2024.

There were no differences in rates of children not on track in any domain or in individual domains in 2020 compared to 2019, but rates were higher in all subsequent years (Table 1).

**Table 1.** Mean and 95% confidence intervals of absolute and relative differences in later years compared to 2019, obtained from 1000 bootstrap simulations.

|  | Any domain | Communication | Gross Motor | Fine Motor | Problem Solving | Personal Social Skills |
| --- | --- | --- | --- | --- | --- | --- |
| Intercept | **17.8 \*\*\*** | **10.7 \*\*\*** | **4.1 \*\*** | **3.4 \*** | **3.1 \*\*** | **5.7 \*\*\*** |
|  | **(10.4 – 30.7)** | **(5.7 – 20.1)** | **(1.6 – 10.5)** | **(1.1 – 10.2)** | **(1.4 – 7.0)** | **(2.7 – 12.2)** |
| Effect of Year (Absolute differences) [Ref: 2019] |  |  |  |  |  |  |
| 2020 | 0.7<br>(-0.3 to 1.9) | 0.7<br>(-0.2 to 1.7) | -0.3<br>(-0.9 to 0.4) | -0.2<br>(-0.8 to 0.5) | 0.4<br>(-0.3 to 0.9) | 0.2<br>(-0.5 to 0.9) |
| 2021 | **1.4 \***<br>**(0.2 to 2.6)** | **1.6 \*\*\***<br>**(0.5 to 2.7)** | 0<br>(-0.7 to 0.7) | 0.1<br>(-0.7 to 0.8) | **0.7 \***<br>**(0.0 to 1.3)** | **0.8 \*\***<br>**(0.1 to 1.5)** |
| 2022 | **3.7 \*\*\***<br>**(2.5 to 4.8)** | **4.1 \*\*\***<br>**(3.1 to 5.1)** | 0.4<br>(-0.3 to 1) | **0.9 \***<br>**(0.1 to 1.6)** | **2.1 \*\*\***<br>**(1.4 to 2.7)** | **2.6 \*\*\***<br>**(1.8 to 3.3)** |
| 2023 | **4.9 \*\*\***<br>**(3.6 to 6.3)** | **4.3 \*\*\***<br>**(3.4 to 5.2)** | **0.9 \*\***<br>**(0.2 to 1.6)** | **1.4 \*\*\***<br>**(0.6 to 2)** | **2.9 \*\*\***<br>**(2.2 to 3.5)** | **3.3 \*\*\***<br>**(2.5 to 4)** |
| 2024 | **4.2 \*\*\***<br>**(2.9 to 5.5)** | **3.6 \*\*\***<br>**(2.5 to 4.6)** | 0.5<br>(-0.2 to 1.2) | **1.0 \*\*\***<br>**(0.2 to 1.7)** | **2.4 \*\*\***<br>**(1.7 to 3.1)** | **2.5 \*\*\***<br>**(1.8 to 3.3)** |
| Effect of Year (Relative differences) [Ref: 2019] |  |  |  |  |  |  |
| 2020 | 1.05<br>(0.98 to 1.14) | 1.08<br>(0.98 to 1.2) | 0.94<br>(0.84 to 1.08) | 0.95<br>(0.82 to 1.12) | 1.08<br>(0.95 to 1.22) | 1.03<br>(0.92 to 1.17) |
| 2021 | **1.1 \***<br>**(1.01 to 1.19)** | **1.18 \*\*\***<br>**(1.06 to 1.33)** | 1<br>(0.86 to 1.15) | 1.01<br>(0.85 to 1.22) | **1.15 \***<br>**(0.99 to 1.31)** | **1.15 \*\***<br>**(1.01 to 1.3)** |
| 2022 | **1.26 \*\*\***<br>**(1.16 to 1.35)** | **1.46 \*\*\***<br>**(1.33 to 1.6)** | 1.08<br>(0.94 to 1.24) | **1.21 \***<br>**(1.03 to 1.41)** | **1.46 \*\*\***<br>**(1.29 to 1.64)** | **1.47 \*\*\***<br>**(1.31 to 1.65)** |
| 2023 | **1.35 \*\*\***<br>**(1.24 to 1.46)** | **1.48 \*\*\***<br>**(1.35 to 1.61)** | **1.19 \*\***<br>**(1.04 to 1.35)** | **1.32 \*\*\***<br>**(1.12 to 1.53)** | **1.63 \*\*\***<br>**(1.44 to 1.81)** | **1.6 \*\*\***<br>**(1.43 to 1.78)** |
| 2024 | **1.3 \*\*\***<br>**(1.19 to 1.4)** | **1.39 \*\*\***<br>**(1.26 to 1.53)** | 1.1<br>(0.95 to 1.27) | **1.23 \*\***<br>**(1.04 to 1.45)** | **1.53 \*\*\***<br>**(1.35 to 1.72)** | **1.46 \*\*\***<br>**(1.3 to 1.64)** |

95% confidence intervals from 1000 bootstrapped simulations presented
\* $p < 0.05$ , \*\* $p < 0.01$ , \*\*\* $p < 0.001$ , $p$ values based on Satterthwaite approximation for the number of degrees of freedom of parameter estimates

For Any Domain, there were progressively larger differences compared to pre-pandemic in cohorts experiencing a longer or earlier age of exposure to the pandemic (2021: 1.4[0.2; 2.6]; 2022: 3.7[2.5; 4.8], 2023: 4.9[3.6; 6.3]). Rates remained higher in 2024 (4.2[2.9; 5.5]) who did not experience the pandemic. The largest absolute increase, in 2023, was equivalent to a 35%[24%; 46%] relative increase.

For Communication, Problem Solving and Personal Social Skills, absolute (Range: 0.7 to 1.6) and relative (Range: 15% to 18%) increases compared to 2019 were small in 2021. Increases were similar in 2022 and 2023, when the largest absolute increase was observed (Communication: 4.3[3.4; 5.2]; Problem solving: 2.9[2.2; 3.5]; Personal Social Skills (3.3[2.5; 4]), equivalent to a 48% to 63% relative increase. The size of increase fell slightly in 2024. For Fine Motor, there were no differences in 2020 and 2021 compared to 2019. Absolute increases were observed in 2022 (0.9[0.1; 1.6]) and 2023 (1.4[0.6; 2]), equivalent to a 21% to 32% relative increase. For Gross Motor, there was a 0.9[0.2; 1.6] absolute increase in 2023, and no differences in other years.

### Socioeconomic inequalities in early childhood development

On average between 2019 and 2024, the rate of children not on track in any domain was higher in LAs in the least advantaged quintile (Rate per 100: 21.3[19.5; 23.0]) compared to most advantaged quintile (16.2[14.7; 17.8]) (Figure 2 and Table S8). The difference was largest in 2022 (6.4[2.9; 9.9]) (Table S9). There was a high level of overlap in the confidence intervals for outcomes within each quintile (Figure S9).

**Figure 2.**
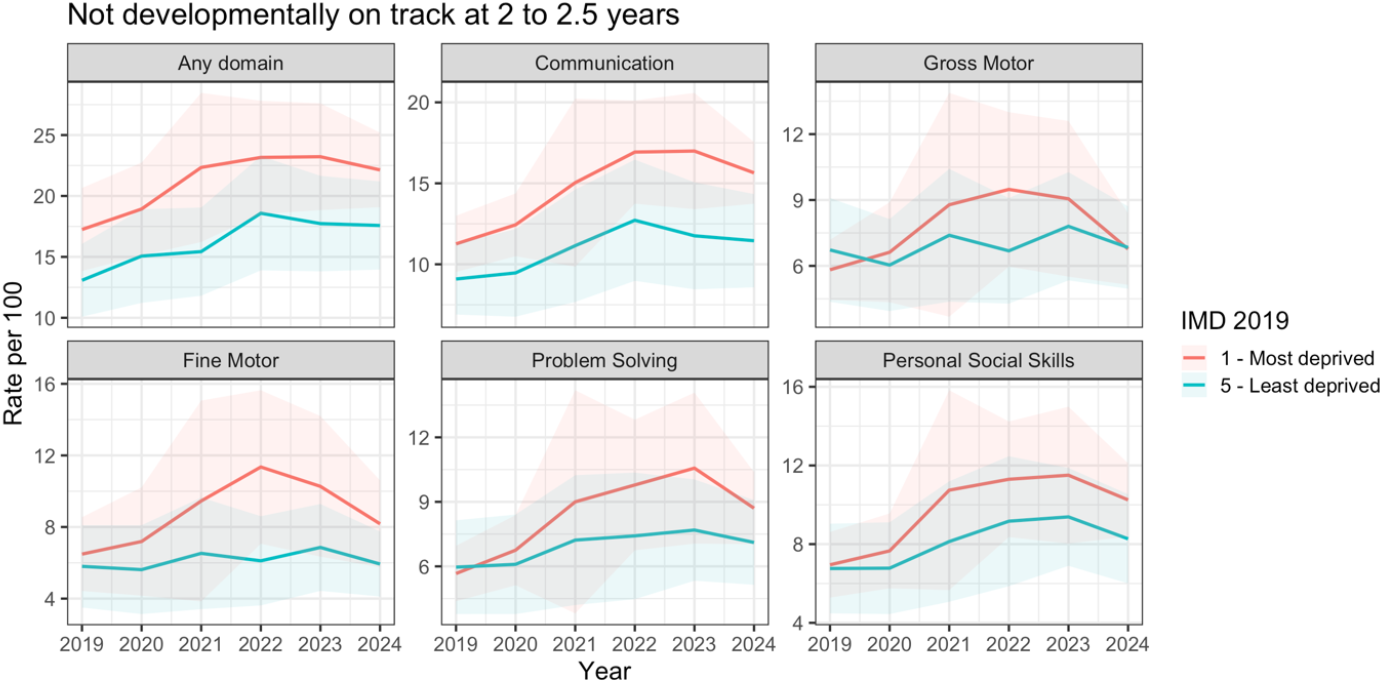
Time trends in the means with 95% confidence intervals for local authorities in the most deprived 20% of Index of Multiple Deprivation Quintile in England, and local authorities in the least deprived quintile.

We did not find evidence of interaction effects of year by area-level socioeconomic conditions (Table S10 and S11). We found evidence for absolute and relative inequalities in rates of children not on track for all outcomes, except Gross Motor, based on Bootstrapped confidence intervals (Table 2, Table S12). For the rate per 100 children not on track in any domain, the Slope Index of Inequality (SII) indicated that the most deprived areas had a 2.9[1.8; 3.9] higher rate, compared to the least deprived areas. The SII was largest for Communication (2.8[1.8; 3.9]). The RII for Any domain was 23%[14%; 32%], and largest for Communication (36%[25%; 49%]) and Fine motor (40%[19%; 62%]), although confidence intervals were wider for Fine Motor.

**Table 2.** Estimates (Mean(95%CI)) of the Slope and Relative Index of Inequality using 1000 bootstrapped samples.

| Index | Any domain | Communication | Gross Motor | Fine Motor | Problem Solving | Personal Social Skills |
| --- | --- | --- | --- | --- | --- | --- |
| SII | 2.9<br>(1.8 to 3.9) | 2.8<br>(2 to 3.6) | 0.2<br>(-0.5 to 0.7) | 1.5<br>(0.7 to 2.1) | 1.1<br>(0.5 to 1.6) | 1<br>(0.4 to 1.5) |
| RII | 1.23<br>(1.14 to 1.32) | 1.36<br>(1.25 to 1.49) | 1.03<br>(0.9 to 1.16) | 1.40<br>(1.19 to 1.62) | 1.27<br>(1.12 to 1.42) | 1.19<br>(1.07 to 1.32) |

Model adjusted for Year, ASQ coverage and ethnic composition

### Sensitivity analyses

We re-ran models using linear and quadratic year, capturing only the continuous effect of year (instead of year-specific COVID-19 effects), to increase power in assessing Year x SECs interactions. We also did not find evidence of interaction effects in these models (Table S13 and S14). To test if findings were robust to data quality, we re-ran models excluding outcome outliers (Appendix S1, Table S15) and found little change in the parameter estimates (Table S16).

### Service-use and economic costs

In 2023, a 5 per 100 increase in the rate of children not on track in any domain could mean 32,000 more children require child health services nationally, using a population estimate of 640,000 2-year-olds.^35^ Worse ECD outcomes in 2023 could incur an additional £60 million annually in early intervention and educational costs, and parental employment losses, and potentially enormous costs due to reduced workforce participation or economic productivity in adulthood (Table 3)

**Table 3.** Summary of cost estimates based on the 2023 increase in children not on track in development, compared to pre-pandemic.

| Cost Category | Cost assumptions (Source) | Cost incurred* |
| --- | --- | --- |
| **Contemporary costs** |  |  |
| Increased early intervention (speech and language therapy) | £2,000 per affected child annually (NHS England, 2023)<sup>36</sup> | £6.4 million annually |
| Increased demand for Special Education Needs support | £10,000 per affected child annually (Education Policy Institute, 2020)<sup>37</sup> | £32 million annually |
| Parental employment losses | Average employment loss of £7,500 per affected child annually (OECD, 2021)<sup>38</sup> | £24 million annually |
| **Predicted future costs** |  |  |
| Lower workforce participation resulting from lower educational attainment | £100,000 reduction in lifetime earnings per child who have lower educational attainment (UK Department for Education, 2021)<sup>39</sup> | £320 million in lifetime earnings loss |
| Reduced economic productivity | 10% reduction in productivity amongst children affected (Penn Wharton Budget Model, 2021)<sup>40</sup> | £1 billion fall in UK GDP<sup>^</sup> |

\*Costs incurred are calculated based on published costs, the 2023 estimate that 5 per 100 more children fail the ASQ-3 developmental screen at 2-2.5 years, and that 10% of children failing the early developmental screen require additional services/greater parental support, and experience lower educational attainment/workforce participation and reduced economic productivity in adulthood (ie., 90% of children affected by COVID-19 catch up in their development over time).<sup>41,42</sup>
<sup>^</sup>UK GDP was USD3.38 trillion (approximately £2.6 trillion) in 2023.<sup>43</sup>

## Discussion

Drawing on English national surveillance data, our study provides timely public health intelligence on recent trends and socioeconomic inequalities in ECD, as early as 2 to 2.5 years. Cohorts assessed during or after the pandemic had greater rates of children not developmentally on track at 2 to 2.5 years, compared to 2019, the unexposed cohort. The greatest increases were amongst children exposed to the pandemic from birth (approximately 5 per 100 increase in 2023, similar increase in 2022), with smaller increases amongst children exposed from 1 year of age (2021) and no differences in children who experienced the shortest exposure (2020). Notably, children born after the pandemic (2024) continued to experience worse outcomes in almost all domains. Stark area-level socioeconomic inequalities, which did not change during this period, were observed in all developmental domains except Gross Motor. The Communication domain had the largest socioeconomic inequalities and greatest pandemic-related increases.

While acknowledging some data quality limitations, we demonstrate the usefulness of a novel dataset in England to monitor the impact of national shocks on ECD. An important limitation is the underestimation of inequalities when data is aggregated over the population in an area.^44^ Studies using individual-level data have found a stronger adverse impact of the pandemic on various domains of development amongst children from low versus high socioeconomic status.^16,19,21,45^ Individual-level data will be best placed to investigate whether the impact of early years exposure to the COVID-19 pandemic may be sustained longitudinally into school age. In the UK, on the ground efforts are needed, including training and increasing staff capacity to contribute to data reporting, to improve data completeness of individual-level data on ASQ-3 in the Community Services Dataset.

Our findings, showing that cohorts exposed to COVID-19 experienced worse ECD outcomes in almost all domains, are consistent with large national studies assessing ECD at around 2 years of age.^46,47^ Our findings support previous work showing that children born during the pandemic, who experienced a longer exposure to the pandemic experienced the worst outcomes.^47^ Previous work has also found socioeconomic inequalities in ECD in all domains before and during the pandemic.^18,19,47^

Differences in methodology and sample may explain differences between our findings and the wider literature. A relatively small study comparing COVID-19 exposed cohort of 2-year-olds to a historical cohort only found differences in the communication domain.^15^ In studies examining ECD in children younger than 1.5 years, or in children of various ages over 0 to 5, experience of COVID-19 on worse development has been inconsistently reported.^12,13,15,17,47– 50^ Age of assessment may be important because some developmental delays may only be identified later.^51^ Domain-level effects may differ by age of exposure to the COVID-19 pandemic. Similar to us, another study reported gross motor delays in younger children exposed to the pandemic, but not in older children,^47^ and exposure to COVID-19 around 2-years-old affected only the communication and social interaction domain at 3-years-old.^16^ Communication may be the most susceptible, as its development relies on the social environment which was severely restricted by the pandemic. This likely explains why communication delays were the most consistently observed across studies, despite methodological differences.

The cost of COVID-19 on and inequalities in children’s early development is likely to be enormous and far-reaching. Implications on the economy, society and health services require immediate attention in pandemic recovery efforts. In 2023 alone, 32,000 more children required further paediatric assessment, adding to already alarming waitlists.^52,53^ The extent of national inequality (3 per 100 absolute difference between the worst and least deprived areas) appears comparable to around two-thirds the adverse developmental impact over the period of the pandemic. An unequal start in life could hinder children from reaching their developmental potential. Spending on late intervention services has risen significantly in the UK post-pandemic, but only small rises seen in early intervention spending, after a decade of cuts to early intervention spending.^54^ Our findings emphasise the need for immediate investment in early intervention, which could include an additional £6.4m annually in speech and language therapy services, and an additional £32m annually in the education sector to support the increase in children with special educational needs. Preventative early intervention can support children to reach their full potential, reduce more costly late intervention services and yield long-term economic benefits through improved workforce participation, higher tax revenues, as well as reduced public expenditure on welfare and criminal justice services.

The wide pre-existing socioeconomic inequalities in ECD prior to the pandemic, alongside worse ECD outcomes since the pandemic, does not bode well for the UK government’s school readiness targets. Children from socioeconomically disadvantaged backgrounds are the most at risk falling behind their peers at school entry. Meeting school readiness targets will require a strong policy focus on inequality reduction, including addressing child poverty, inequalities in the home learning environment, and access to early childhood education. The early childhood education and care sector is a key sector to alleviate inequalities in ECD.^55^ Efforts to tackle the post-pandemic challenges faced by the sector, such as reduced attendance and workforce issues, are urgently needed.^56^

To conclude, we report worsening ECD outcomes at 2 to 2.5 years in England, alongside stark socioeconomic inequalities, during and after the pandemic. All domains were affected, with the most pronounced impact on communicative development. The cost of the pandemic’s impact on ECD is likely to be enormous. Ensuring children have a fair start in life requires investment in the early years. National ECD data can enable monitoring outcomes post-pandemic and progress in closing inequality gaps.

## Data Availability

Data on Ages and Stages Questionnaire aggregated to local authority level was openly available on Fingertips: https://fingertips.phe.org.uk/profile/child-health-profiles/data#page/7/gid/1938133402/pat/159/par/K02000001/ati/15/are/E92000001/iid/93436/age/241/sex/4/cat/-1/ctp/-1/yrr/1/cid/4/tbm/1/page-options/ine-vo-1_ine-yo-1:2020:-1:-1_ine-pt-0_ine-ct-113

## Acknowledgements

YWC is part-funded by an NIHR Research Professorship (NIHR302438) awarded to DTR. CM was funded by an NIHR Undergraduate Internship Programme (NIHR304390), awarded to YWC. DTR and DB are funded by the NIHR School for Public Health Research (grant reference number NIHR204000). DTR is also funded on an NIHR Research Professorship (NIHR302438). LM is part funded by the NIHR Applied Research Collaboration Greater Manchester (NIHR200174). The views expressed are those of the authors and not necessarily those of the NIHR or the Department of Health and Social Care.

## Notes

**Conflicts of interests:** The authors declare no conflict of interests.

### Competing Interest Statement

The authors have declared no competing interest.

### Funding Statement

The study received funding from the National Institute for Health and Care Research (NIHR)

### Summary of Updates

Title page updated - author affiliation

