## Appendices for "Time trends and area-level socioeconomic inequalities in early childhood development at 2 to 2.5 years old in England between 2019 and 2024"

**S1. Supplemental Methods**
**S2. Directed Acyclic Graphs**
**S3. Bias due to poor ASQ coverage**
**S4. Supplemental Tables S1 to S15**
**S5. Supplemental Figures S1 to S8**

### Appendix S1. Supplemental methods

#### *Excluded data due to local authority boundaries*

Data from Cumberland and Westmorland and Furness were excluded, as only available for 2023/24 due to new local authority boundaries and there was no data on IMD 2019. Data from City of London was combined with Hackney. Data from Isles of Scilly was combined with Cornwall.

#### *Data excluded based on preliminary analyses*

In preliminary analyses, we described data quality and coverage over time. We obtained the numbers and percentage of local authorities submitting information on the ASQ, distribution in coverage of mandated contacts, and correlation using Kendall rank correlation coefficient between ASQ coverage and IDACI Average Scores within each year. We obtained sample statistics (e.g. mean, standard deviation, skew, kurtosis) and visualised the spread of data using boxplots, histograms and q-q plots of each early child developmental outcome over all local authorities and years. For all analyses with IDACI Average Scores, data for West Northamptonshire and North Northamptonshire were combined to provide data for Northamptonshire, reflecting local authority boundaries in 2019.

We examined the most extreme 1% of data points with the highest rate of children not developmentally on track in any domain. For example, the highest three rates were 95.8 per 100 children, 89.1 per 100 children and 77.1 per 100 children. We excluded data from Barnet (2023), Ealing (2023, 2024), Richmond upon Thames (2023), Stoke-on-Trent (2018) and Brent (2020). We decided to exclude 2023 data for Barnet (over 800 per 1000 children did not achieve a good level of development in each of the five domain, an anomaly for that year), 2023 and 2024 data from Ealing (Over 75 per 100 children did not achieve a good level of development in communication, compared to less than 10 per 100 children in all other domains, and anomalous from previous years), 2023 data for Richmond-Upon Thames (over 60 per 100 children did not achieve a good level of development in communication compared to less than 50 per children in other domains, anomalous from all other years), 2018 for Stoke-on-Trent and 2020 for Brent (anomalously high rates of children who did not achieve a good level of development in first year of available data, compared to all the other years).

On the lower extreme, we excluded data from Torbay in 2020 and 2024, due to rates of 0 for personal social skills.

We tabulated the number of LAs with outliers above the upper IQR limit (see Table S4) and conducted sensitivity analyses excluding these outliers.

#### *Model equations*

Let:

$Dev_{it}$ : Rate of children who do not achieve a good level of development, expressed in rate per 1000

$Year_t$: Indicator variable with six levels, with 2018/19 as the reference year, 2019/20, 2020/21, 2021/22, 2022/23, and 2023/24

$IDACI_i$: Population-weighted rank (scaled to 0 to 1) of Index of Multiple Deprivation 2019, Income Deprivation Affecting Children Index for each *i* representing *i-th* local authority

$Review_{it}$: Percentage of eligible population receiving an ASQ review in each year for each *i* representing *i-th* local authority

$Emp_{it}$: Percentage of households with children aged 0 to 15 years who are employed in each year, for each *i* representing *i-th* local authority

$Ethnicity_i$: Percentage of children 0 to 15 years of White ethnicity based on 2021 census, for each *i* representing *i-th* local authority

$\gamma_i$: amount of variation in $\beta_{0i}$ for the *i-th* local authority, assumed to follow a normal distribution $N(0, \sigma^2)$

$\varepsilon_{it}$: residual error

Model equation for the effect of Year

$$Dev_{it} = \beta_{0i} + \beta_1 Year_t + \beta_2 Review_{it} + \beta_3 Emp_{it} + \beta_4 Ethnicity_i + \beta_5 IDACI_i + \gamma_i + \varepsilon_{it}$$

Model equation for the effect of IDACI

$$Dev_{it} = \beta_{0i} + \beta_1 Year_t + \beta_2 IDACI_i + \beta_3 IDACI_i Year_t + \beta_4 Review_{it} + \beta_5 Ethnicity_i + \gamma_i + \varepsilon_{it}$$

***Service-use and economic costs***

We use English midyear population estimates of approximately 640,000 2-year-olds to calculate the increase in number of children requiring further paediatric assessment.[1] We assumed that all children falling behind based on the ASQ-3 developmental screen require further paediatric assessment. For both contemporaneous and future costs, we make a conservative assumption that 10% of children failing the screen at 2 to 2.5 years require early interventions and may experience sustained long-term consequences. In other words, we assume that 90% of the additional 5 per 100 children at some point ‘recover’ to the pre-COVID-19 trajectories.[2,3] Table S2 summarises how the costs were calculated.

For contemporary costs, we expect that some children experiencing developmental delays require early intervention (we focus on speech and language therapy since communication delays are the most prevalent), special educational needs support when they enter school, and additional parental support which may result in parents reducing employment to fulfil childcare duties.

Longitudinal studies in the UK have shown that children showing early developmental delays are less likely to achieve five GCSEs at grade C and above.[4] Further, recent UK Department for Education estimates that failing to achieve this educational threshold is associated with £96,000 in lifetime earnings.[5] Based on this estimates, we consider future costs on workforce participation resulting from lower educational attainment. In addition, the Penn Wharton Budget Model (2021) suggests that investments in early childhood development yield long-term economic returns through increased productivity and tax revenue. Therefore, potential cascading effects of developmental delays on adulthood workforce productivity and affect unemployment rates, may contribute to economic losses.

5. Hodge L, Little A, Weldon M. *GCSE Attainment and Lifetime Earnings*. https://assets.publishing.service.gov.uk/media/60c36f0cd3bf7f4bd11a2326/GCSE_Attainment_and_Lifetime_Earnings_PDF3A.pdf

### Appendix S2. Directed Acyclic Graphs

We were interested in using Year to proxy the effect of COVID-19 shocks. To analyse the effect of Year on the Outcome, we controlled for factors affecting review (biasing pathway) by conditioning percent white ethnicity, income deprivation, and ASQ coverage as a proxy of any residual factors affecting review. We also controlled for improving employment trends in general, that was not resulting from COVID-19 shocks.

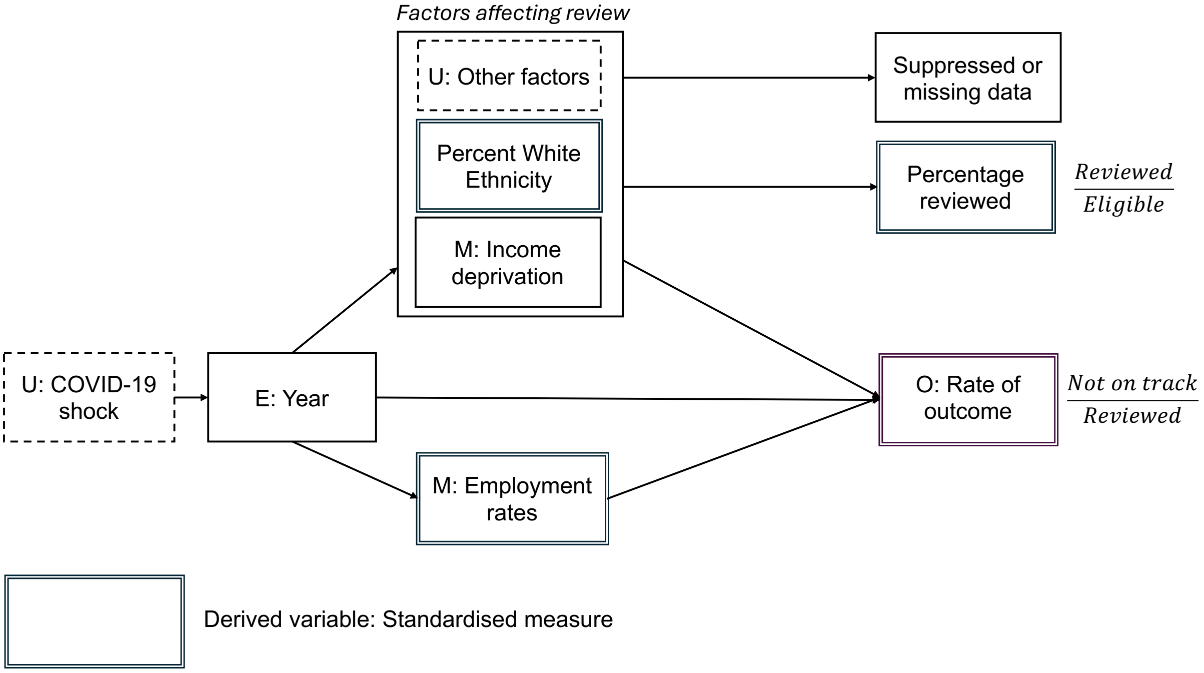


To analyse the effect of Income deprivation on the outcome, we controlled for Year and Percent White ethnicity as confounders. We controlled for other factors affecting review which could also confound the relationship, by controlling for ASQ coverage. We did not control for employment rates as were interested in the full extent which income deprivation affects the outcome, including when income deprivation occurs in the context of unemployment.

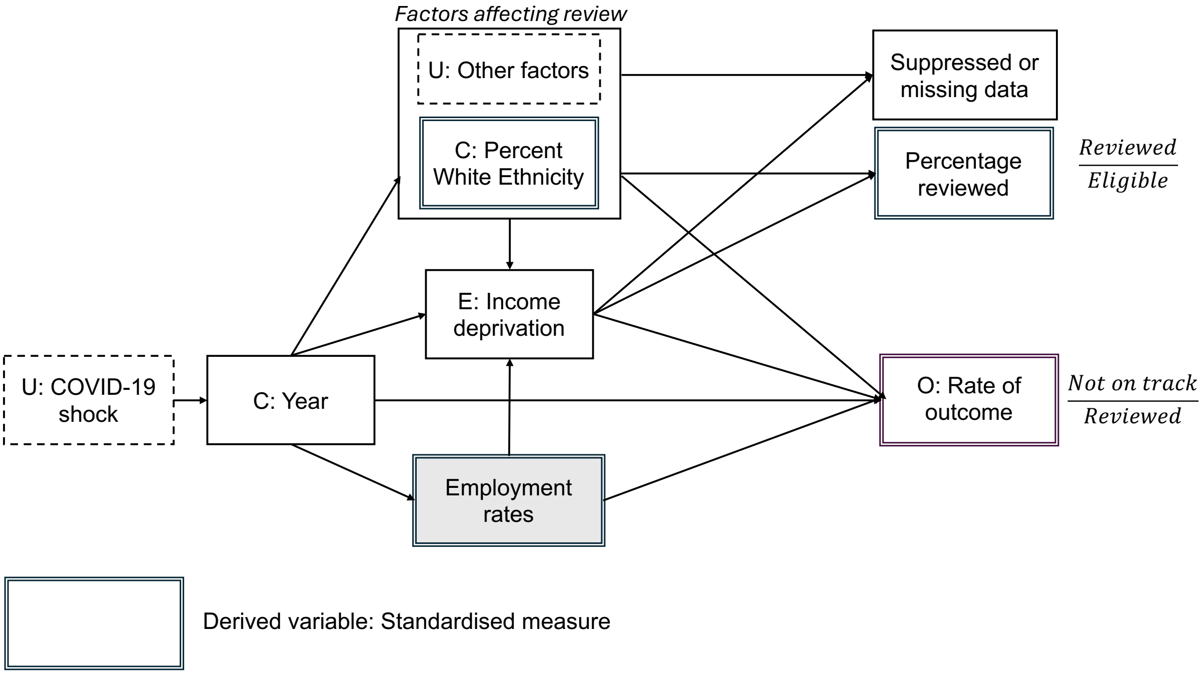


Figure. Directed Acyclic Graph showing the relationship of interest: income deprivation on the rate of children not developmentally on track.

### Appendix S3. Bias due to poor ASQ coverage

Children of particular characteristics (e.g. non-white ethnicity are less likely to be reviewed). In addition these characteristics may be associated with poorer early childhood outcomes. This leads to selection bias as we are only able to analyse the outcome amongst children who have been reviewed. As such, amongst local authorities with lower coverage, we expect early childhood developmental outcomes to be better as a result of missing children who have poorer outcomes.

Limiting the analysis only to local authorities with non-missing data and high coverage, however, may also lead to selection bias. This could result in excluding local authorities with high deprivation or higher percentage of non-white ethnicity – factors which are potentially associated with lower coverage, and which are associated with poorer outcomes.

Most local authorities have coverage above 90% (Table S6 and Figure SX). As such we decided to analyse data in local authorities with adequate coverage, defined as 75% or greater coverage. To address potential bias due to variation in coverage, in analyses of Year, we conditioned on ASQ coverage, percent White ethnicity and Income Deprivation, so that we analysed the relationship of interest amongst local authorities with the same level of coverage (ie., conditioning for factors affecting coverage). In analyses of Income Deprivation we conditioned on ASQ coverage, percent White ethnicity and Year.

### Appendix S4. Supplemental Tables

Table S1. Descriptive statistics of ECD outcomes

| **Sample statistics** | **All five domains** | **Communication** | **Gross Motor** | **Fine Motor** | **Problem Solving** | **Personal Social Skills** |
|---|---|---|---|---|---|---|
| **n** | 854 | 860 | 858 | 858 | 857 | 858 |
| **mean** | 19.0 | 13.7 | 7.5 | 7.6 | 8.1 | 9.2 |
| **sd** | 10.4 | 10.6 | 9.1 | 9.7 | 9.2 | 9.5 |
| **se** | 0.4 | 0.4 | 0.3 | 0.3 | 0.3 | 0.3 |
| **median** | 16.1 | 11.0 | 5.0 | 4.5 | 5.7 | 6.8 |
| **IQR** | 9.8 | 7.2 | 4.7 | 5.9 | 4.8 | 5.8 |
| **min** | 2.5 | 0.4 | 0.2 | 0.2 | 0.5 | 0.0 |
| **max** | 95.9 | 99.2 | 99.2 | 99.2 | 99.2 | 99.2 |
| **skew** | 2.3 | 4.1 | 5.8 | 4.9 | 5.6 | 5.5 |
| **kurtosis** | 9.0 | 24.1 | 46.6 | 35.6 | 44.3 | 42.2 |

Table S2. Summary of how costs were calculated

| **Cost Category** | **Cost assumptions (Source)** | **Cost incurred** |
|---|---|---|
| Contemporary costs | | |
| Increased early intervention (speech and language therapy) | £2,000 per affected child annually (NHS England, 2023)[6] | (640,000*0.05*0.1*£2,000) |
| Increased demand for Special Education Needs support | £10,000 per affected child annually (Education Policy Institute, 2020)[7] | (640,000*0.05*0.1*£10,000) |
| Parental Employment Losses | Average employment loss of £7,500 per affected child annually (OECD, 2021)[8] | (640,000*0.05*0.1*£7,500) |
| Predicted future costs | | |
| Lower Educational Attainment and Workforce Participation | £100,000 reduction in lifetime earnings per child who have lower educational attainment (UK Department for Education, 2021)[5] | (640,000*0.05*0.1*£100,000) |
| Reduced Economic Productivity | 10% reduction in productivity amongst children affected (Penn Wharton Budget Model, 2021)[9] | 10%*UK GDP^ |

^USD 3.38 trillion in 2023[10]

Table S3. Number (%) of local authorities with published data on the ASQ-3 2018 to 2024

| Year | All five domains | Communication | Gross Motor | Fine Motor | Problem Solving | Personal social skills |
|---|---|---|---|---|---|---|
| 2018 | 74 (49.7%) | 75 (50.3%) | 75 (50.3%) | 75 (50.3%) | 75 (50.3%) | 75 (50.3%) |
| 2019 | 113 (75.8%) | 116 (77.9%) | 115 (77.2%) | 115 (77.2%) | 115 (77.2%) | 115 (77.2%) |
| 2020 | 129 (86.6%) | 127 (85.2%) | 127 (85.2%) | 127 (85.2%) | 127 (85.2%) | 127 (85.2%) |
| 2021 | 134 (89.9%) | 135 (90.6%) | 134 (89.9%) | 134 (89.9%) | 134 (89.9%) | 134 (89.9%) |
| 2022 | 139 (93.3%) | 139 (93.3%) | 139 (93.3%) | 139 (93.3%) | 138 (92.6%) | 139 (93.3%) |
| 2023 | 133 (89.3%) | 133 (89.3%) | 133 (89.3%) | 133 (89.3%) | 133 (89.3%) | 133 (89.3%) |
| 2024 | 138 (92.6%) | 138 (92.6%) | 138 (92.6%) | 138 (92.6%) | 138 (92.6%) | 138 (92.6%) |

Table S4. Correlation between ASQ Coverage and IDACI Average Score

| Year | Kendall tau | p |
|---|---:|---:|
| 2018 | 0.01 | 0.816 |
| 2019 | -0.12 | 0.031 |
| 2020 | -0.08 | 0.148 |
| 2021 | -0.06 | 0.292 |
| 2022 | -0.09 | 0.132 |
| 2023 | -0.05 | 0.427 |
| 2024 | -0.01 | 0.914 |

Table S5. ASQ Coverage amongst local authorities reporting data on coverage

| Year | 0% ≤ x < 25% | 25% ≤ x < 50% | 50% ≤ x < 75% | 75% ≤ x ≤ 100% | Total |
|---|---|---|---|---|---|
| 2018 | 18 | 0 | 7 | 113 | 138 |
| 2019 | 13 | 0 | 4 | 127 | 144 |
| 2020 | 4 | 0 | 4 | 133 | 141 |
| 2021 | 15 | 6 | 9 | 108 | 138 |
| 2022 | 16 | 1 | 9 | 118 | 144 |
| 2023 | 14 | 1 | 5 | 113 | 133 |
| 2024 | 14 | 0 | 5 | 125 | 144 |

Table S6. Analysed data – number (%) of local authorities with published data on ASQ and published data on coverage more than 75%, and excluding extreme outliers, and data on 2019 IDACI

| **Year** | **All five domains** | **Communication** | **Gross Motor** | **Fine Motor** | **Problem Solving** | **Personal social skills** |
|---|---|---|---|---|---|---|
| 2018 | NA | NA | NA | NA | NA | NA |
| 2019 | 103 (69.1%) | 106 (71.1%) | 106 (71.1%) | 106 (71.1%) | 106 (71.1%) | 106 (71.1%) |
| 2020 | 119 (79.9%) | 119 (79.9%) | 119 (79.9%) | 119 (79.9%) | 119 (79.9%) | 119 (79.9%) |
| 2021 | 101 (67.8%) | 101 (67.8%) | 101 (67.8%) | 101 (67.8%) | 101 (67.8%) | 101 (67.8%) |
| 2022 | 112 (75.2%) | 112 (75.2%) | 112 (75.2%) | 112 (75.2%) | 111 (74.5%) | 112 (75.2%) |
| 2023 | 106 (71.1%) | 106 (71.1%) | 106 (71.1%) | 106 (71.1%) | 106 (71.1%) | 106 (71.1%) |
| 2024 | 118 (79.2%) | 118 (79.2%) | 118 (79.2%) | 118 (79.2%) | 118 (79.2%) | 118 (79.2%) |

Table S7. Mean (95%CI) rate per 100 children not developmentally on track 2019 to 2024

| Year | All five domains | Communication | Gross Motor | Fine Motor | Problem Solving | Personal social skills |
|---|---|---|---|---|---|---|
| 2019 | 15.6 (14.2 to 16.9) | 10.0 (9.1 to 10.8) | 6.1 (5.2 to 6.9) | 5.9 (4.9 to 6.9) | 5.5 (4.7 to 6.3) | 6.6 (5.7 to 7.5) |
| 2020 | 16.4 (14.9 to 17.9) | 11.0 (9.9 to 12.1) | 5.8 (4.9 to 6.6) | 5.6 (4.6 to 6.6) | 5.9 (5 to 6.8) | 6.6 (5.8 to 7.5) |
| 2021 | 17.6 (15.6 to 19.6) | 12.7 (10.9 to 14.4) | 7 (5.5 to 8.6) | 6.8 (5.1 to 8.4) | 7 (5.4 to 8.5) | 8.2 (6.6 to 9.7) |
| 2022 | 19.7 (18 to 21.5) | 14.5 (13.1 to 15.9) | 7 (5.9 to 8.1) | 7.3 (5.9 to 8.7) | 8 (6.8 to 9.2) | 9.5 (8.3 to 10.7) |
| 2023 | 20.6 (18.9 to 22.4) | 14.7 (13.1 to 16.2) | 7.8 (6.5 to 9.1) | 7.8 (6.4 to 9.2) | 8.8 (7.5 to 10) | 10.1 (8.8 to 11.5) |
| 2024 | 19.8 (18.2 to 21.4) | 13.7 (12.5 to 14.9) | 6.6 (5.9 to 7.4) | 6.8 (5.8 to 7.8) | 7.9 (7 to 8.7) | 9.0 (8.1 to 9.9) |

Table S8. Mean (95% CI) of the time averaged rate per 100 children not developmentally on track, by IMD quintile

| IDACI Quintile | All five domains | Communication | Gross Motor | Fine Motor | Problem Solving | Personal social skills |
|---|---|---|---|---|---|---|
| 1 - Most deprived | 21.3 (19.5 to 23) | 14.8 (13.5 to 16.1) | 7.8 (6.5 to 9) | 8.8 (7.3 to 10.4) | 8.5 (7.2 to 9.7) | 9.8 (8.6 to 11) |
| 2 | 17.4 (16.1 to 18.6) | 12.6 (11.7 to 13.5) | 5.6 (4.8 to 6.3) | 5.9 (4.9 to 6.8) | 6.6 (5.8 to 7.3) | 7.4 (6.6 to 8.2) |
| 3 | 16.9 (15.5 to 18.3) | 11.8 (10.6 to 13.1) | 5.5 (4.6 to 6.4) | 5.7 (4.6 to 6.9) | 6.5 (5.5 to 7.4) | 7.4 (6.4 to 8.4) |
| 4 | 19.7 (18.2 to 21.2) | 13.6 (12.2 to 15) | 7.7 (6.7 to 8.6) | 6.8 (5.7 to 7.8) | 7.4 (6.3 to 8.5) | 9 (7.9 to 10.1) |
| 5 - Least deprived | 16.2 (14.7 to 17.8) | 10.9 (9.7 to 12.2) | 6.9 (5.9 to 7.8) | 6.1 (5.1 to 7.1) | 6.9 (5.9 to 7.9) | 8 (7 to 9.1) |

Table S9. Difference (95% CI) in rate per 100 children not developmentally on track (Most deprived compared to least deprived IMD quintile)

| **Year** | **All five domains** | **Communication** | **Gross Motor** | **Fine Motor** | **Problem Solving** | **Personal social skills** |
|---|---|---|---|---|---|---|
| 2019 | 2.2 (2.6 to 1.7) | -0.9 (0.1 to -1.9) | 0.7 (0.9 to 0.4) | -0.3 (0.6 to -1.2) | 0.2 (0.8 to -0.4) | 4.2 (3.7 to 4.6) |
| 2020 | 3 (3.7 to 2.2) | 0.6 (0.4 to 0.8) | 1.6 (1 to 2.1) | 0.7 (1.3 to 0) | 0.9 (1.3 to 0.5) | 3.9 (3.9 to 3.8) |
| 2021 | 3.9 (2.2 to 5.6) | 1.4 (-0.7 to 3.5) | 2.9 (0.4 to 5.4) | 1.8 (-0.4 to 4) | 2.6 (0.6 to 4.6) | 6.9 (4.4 to 9.4) |
| 2022 | 4.2 (4.8 to 3.7) | 2.8 (1.7 to 3.9) | 5.2 (3.4 to 7) | 2.4 (2.3 to 2.5) | 2.1 (2.5 to 1.8) | 4.6 (4.6 to 4.5) |
| 2023 | 5.2 (4.9 to 5.5) | 1.3 (0.2 to 2.3) | 3.4 (1.9 to 4.9) | 2.9 (1.7 to 4) | 2.1 (1.1 to 3.1) | 5.5 (5 to 6) |
| 2024 | 4.2 (5.2 to 3.2) | -0.1 (0.2 to -0.3) | 2.2 (1.6 to 2.9) | 1.6 (1.9 to 1.3) | 2 (2.4 to 1.6) | 4.6 (5.1 to 4) |

Table S10. Model examining effect of IDACIn including interaction effects, controlled for ASQ coverage, ethnicity and year

| | All five domains | Communication | Gross Motor | Fine Motor | Problem Solving | Personal social skills |
|---|---|---|---|---|---|---|
| *Predictors* | *Estimates* | *Estimates* | *Estimates* | *Estimates* | *Estimates* | *Estimates* |
| (Intercept) | 16.47 $^{***}$ | 10.08 $^{***}$ | 3.53 $^{*}$ | 1.99 | 2.27 | 4.34 $^{***}$ |
| | (9.36 – 28.97) | (5.32 – 19.10) | (1.34 – 9.30) | (0.64 – 6.21) | (0.97 – 5.29) | (1.98 – 9.53) |
| Year [2020] | 1.09 | 0.99 | 0.88 | 0.97 | 1.05 | 1.03 |
| | (0.95 – 1.25) | (0.84 – 1.16) | (0.69 – 1.12) | (0.73 – 1.28) | (0.85 – 1.29) | (0.85 – 1.24) |
| Year [2021] | 1.18 $^{*}$ | 1.17 | 1.11 | 1.32 | 1.29 $^{*}$ | 1.29 $^{*}$ |
| | (1.02 – 1.35) | (0.99 – 1.38) | (0.86 – 1.42) | (0.99 – 1.76) | (1.04 – 1.60) | (1.06 – 1.58) |
| Year [2022] | 1.30 $^{***}$ | 1.38 $^{***}$ | 1.01 | 1.19 | 1.34 $^{**}$ | 1.43 $^{***}$ |
| | (1.13 – 1.50) | (1.17 – 1.63) | (0.79 – 1.30) | (0.89 – 1.59) | (1.07 – 1.66) | (1.17 – 1.75) |
| Year [2023] | 1.41 $^{***}$ | 1.37 $^{***}$ | 1.2 | 1.42 $^{*}$ | 1.53 $^{***}$ | 1.65 $^{***}$ |
| | (1.22 – 1.62) | (1.16 – 1.62) | (0.93 – 1.54) | (1.06 – 1.90) | (1.23 – 1.90) | (1.35 – 2.02) |
| Year [2024] | 1.38 $^{***}$ | 1.35 $^{***}$ | 1.18 | 1.37 $^{*}$ | 1.49 $^{***}$ | 1.47 $^{***}$ |
| | (1.21 – 1.59) | (1.15 – 1.58) | (0.93 – 1.50) | (1.04 – 1.82) | (1.21 – 1.83) | (1.21 – 1.78) |
| COV Value | 1 | 1 | 1 | 1.01 | 1.01 $^{*}$ | 1 |
| | (1.00 – 1.01) | (1.00 – 1.01) | (0.99 – 1.01) | (1.00 – 1.02) | (1.00 – 1.02) | (1.00 – 1.01) |
| IDACIn | 1.36 $^{*}$ | 1.29 | 1.06 | 1.70 $^{*}$ | 1.27 | 1.28 |
| | (1.06 – 1.76) | (0.98 – 1.70) | (0.70 – 1.62) | (1.03 – 2.81) | (0.88 – 1.84) | (0.91 – 1.81) |
| white ethnicity percent | 1.00 $^{***}$ | 0.99 $^{***}$ | 1 | 0.99 | 1 | 1.00 $^{*}$ |
| | (0.99 – 1.00) | (0.99 – 1.00) | (1.00 – 1.00) | (0.99 – 1.00) | (0.99 – 1.00) | (0.99 – 1.00) |
| Year [2020] × IDACIn | 0.92 | 1.17 | 1.12 | 0.95 | 1.05 | 0.99 |
| | (0.74 – 1.15) | (0.90 – 1.52) | (0.75 – 1.67) | (0.60 – 1.51) | (0.74 – 1.49) | (0.72 – 1.37) |
| Year [2021] × IDACIn | 0.86 | 0.99 | 0.81 | 0.58 $^{*}$ | 0.78 | 0.78 |
| | (0.68 – 1.09) | (0.76 – 1.30) | (0.54 – 1.22) | (0.36 – 0.93) | (0.54 – 1.11) | (0.56 – 1.09) |
| Year [2022] × IDACIn | 0.91 | 1.06 | 1.11 | 0.96 | 1.12 | 1 |
| | (0.72 – 1.14) | (0.81 – 1.39) | (0.73 – 1.67) | (0.59 – 1.54) | (0.78 – 1.61) | (0.72 – 1.39) |
| Year [2023] × IDACIn | 0.89 | 1.11 | 0.97 | 0.81 | 1.07 | 0.89 |
| | (0.71 – 1.13) | (0.85 – 1.46) | (0.64 – 1.47) | (0.50 – 1.31) | (0.75 – 1.53) | (0.64 – 1.24) |
| Year [2024] × IDACIn | 0.85 | 1.02 | 0.86 | 0.75 | 1 | 0.94 |
| | (0.68 – 1.07) | (0.78 – 1.32) | (0.58 – 1.29) | (0.47 – 1.19) | (0.71 – 1.42) | (0.68 – 1.30) |
| Random Effects | | | | | | |
| $\sigma^2$ | 0.06 | 0.09 | 0.2 | 0.26 | 0.15 | 0.13 |
| $\tau_{00}$ | $0.11_{\text{AreaName}}$ | $0.12_{\text{AreaName}}$ | $0.27_{\text{AreaName}}$ | $0.41_{\text{AreaName}}$ | $0.21_{\text{AreaName}}$ | $0.19_{\text{AreaName}}$ |
| ICC | 0.64 | 0.57 | 0.58 | 0.61 | 0.59 | 0.6 |
| N | $143_{\text{AreaName}}$ | $143_{\text{AreaName}}$ | $143_{\text{AreaName}}$ | $143_{\text{AreaName}}$ | $143_{\text{AreaName}}$ | $143_{\text{AreaName}}$ |
| Observations | 659 | 662 | 662 | 662 | 661 | 662 |
| Marginal $R^2$ / Conditional $R^2$ | 0.140 / 0.694 | 0.197 / 0.657 | 0.016 / 0.584 | 0.059 / 0.631 | 0.114 / 0.634 | 0.122 / 0.645 |
| AIC | 449.24 | 631.619 | 1170.25 | 1370.521 | 988.936 | 895.342 |

* $p<0.05$ ** $p<0.01$ *** $p<0.001$

Table S11. Likelihood Ratio Tests of models including interactions of categorical Year and IDACI, compared to models without interactions

| Model | LogLik | Chi | p | df |
|---|---|---|---|---|
| I_asq | -175.15 | | | |
| Ii_asq | -173.98 | 2.3347 | 0.8012 | 5 |
| I_comm | -268.24 | | | |
| Ii_comm | -267.06 | 2.3646 | 0.7967 | 5 |
| I_gm | -544.22 | | | |
| Ii_gm | -542.22 | 4.0152 | 0.5458 | 5 |
| I_fm | -648.18 | | | |
| Ii_fm | -644.51 | 7.3361 | 0.1968 | 5 |
| I_ps | -452.17 | | | |
| Ii_ps | -449.62 | 5.0987 | 0.404 | 5 |
| I_pss | -403.45 | | | |
| Ii_pss | -401.78 | 3.3398 | 0.6477 | 5 |

Table S12. Model examining effect of IDACIn excluding interaction effects, controlled for ASQ coverage, ethnicity and year

| | **All five domains** | **Communication** | **Gross Motor** | **Fine Motor** | **Problem Solving** | **Personal social skills** |
|---|---|---|---|---|---|---|
| *Predictors* | *Estimates* | *Estimates* | *Estimates* | *Estimates* | *Estimates* | *Estimates* |
| (Intercept) | 17.93 *** | 9.90 *** | 3.78 ** | 2.38 | 2.32 * | 4.59 *** |
| | (10.30 – 31.20) | (5.29 – 18.50) | (1.46 – 9.76) | (0.78 – 7.28) | (1.01 – 5.32) | (2.12 – 9.92) |
| Year [2020] | 1.04 | 1.07 | 0.94 | 0.94 | 1.07 | 1.02 |
| | (0.98 – 1.12) | (0.99 – 1.16) | (0.83 – 1.05) | (0.82 – 1.08) | (0.97 – 1.19) | (0.93 – 1.12) |
| Year [2021] | 1.09 * | 1.17 *** | 0.99 | 1 | 1.13 * | 1.14 * |
| | (1.02 – 1.17) | (1.07 – 1.27) | (0.88 – 1.13) | (0.86 – 1.15) | (1.02 – 1.26) | (1.03 – 1.26) |
| Year [2022] | 1.24 *** | 1.43 *** | 1.07 | 1.17 * | 1.42 *** | 1.43 *** |
| | (1.16 – 1.33) | (1.32 – 1.55) | (0.95 – 1.21) | (1.01 – 1.34) | (1.28 – 1.58) | (1.30 – 1.58) |
| Year [2023] | 1.32 *** | 1.45 *** | 1.18 ** | 1.27 ** | 1.58 *** | 1.55 *** |
| | (1.24 – 1.42) | (1.33 – 1.57) | (1.04 – 1.33) | (1.10 – 1.47) | (1.42 – 1.76) | (1.40 – 1.71) |
| Year [2024] | 1.28 *** | 1.36 *** | 1.09 | 1.18 * | 1.49 *** | 1.42 *** |
| | (1.19 – 1.37) | (1.26 – 1.47) | (0.97 – 1.23) | (1.03 – 1.36) | (1.34 – 1.66) | (1.29 – 1.56) |
| IDACIn | 1.23 * | 1.36 ** | 1.03 | 1.4 | 1.27 | 1.19 |
| | (1.00 – 1.51) | (1.10 – 1.68) | (0.75 – 1.43) | (0.95 – 2.08) | (0.95 – 1.69) | (0.91 – 1.56) |
| COV Value | 1 | 1 | 1 | 1.01 | 1.01 * | 1 |
| | (0.99 – 1.01) | (1.00 – 1.01) | (0.99 – 1.01) | (1.00 – 1.02) | (1.00 – 1.02) | (1.00 – 1.01) |
| white ethnicity percent | 1.00 *** | 0.99 *** | 1 | 0.99 | 1 | 1.00 * |
| | (0.99 – 1.00) | (0.99 – 1.00) | (1.00 – 1.00) | (0.99 – 1.00) | (0.99 – 1.00) | (0.99 – 1.00) |
| **Random Effects** | | | | | | |
| $\sigma^2$ | 0.06 | 0.09 | 0.2 | 0.27 | 0.15 | 0.13 |
| $\tau_{00}$ | 0.11 AreaName | 0.12 AreaName | 0.27 AreaName | 0.41 AreaName | 0.21 AreaName | 0.19 AreaName |
| ICC | 0.64 | 0.57 | 0.57 | 0.6 | 0.58 | 0.59 |
| N | 143 AreaName | 143 AreaName | 143 AreaName | 143 AreaName | 143 AreaName | 143 AreaName |
| Observations | 659 | 662 | 662 | 662 | 661 | 662 |
| Marginal $R^2$ / Conditional $R^2$ | 0.138 / 0.692 | 0.196 / 0.656 | 0.013 / 0.581 | 0.054 / 0.626 | 0.111 / 0.631 | 0.119 / 0.643 |
| AIC | 427.547 | 611.6 | 1156.016 | 1360.992 | 974.366 | 878.239 |

* *p<0.05* ** *p<0.01* *** *p<0.001*

Table S13. Models using continuous Year (linear and quadratic effect, with interaction effect with IDACI)

| | **All five domains** | **Communication** | **Gross Motor** | **Fine Motor** | **Problem Solving** | **Personal social skills** |
|---|---|---|---|---|---|---|
| *Predictors* | *Estimates* | *Estimates* | *Estimates* | *Estimates* | *Estimates* | *Estimates* |
| (Intercept) | 16.47 *** | 9.81 *** | 3.71 ** | 2.13 | 2.24 | 4.39 *** |
| | (9.39 – 28.89) | (5.18 – 18.57) | (1.42 – 9.73) | (0.68 – 6.62) | (0.96 – 5.22) | (2.00 – 9.67) |
| Year | 1.12 ** | 1.15 ** | 1.02 | 1.13 | 1.16 * | 1.22 ** |
| | (1.03 – 1.22) | (1.04 – 1.27) | (0.88 – 1.18) | (0.95 – 1.35) | (1.02 – 1.33) | (1.08 – 1.37) |
| $Year^2$ | 0.99 | 0.99 | 1.01 | 0.99 | 0.99 | 0.98 |
| | (0.98 – 1.01) | (0.97 – 1.01) | (0.98 – 1.03) | (0.96 – 1.02) | (0.96 – 1.01) | (0.96 – 1.00) |
| COV Value | 1 | 1 | 1 | 1.01 | 1.01 * | 1 |
| | (1.00 – 1.01) | (1.00 – 1.01) | (0.99 – 1.01) | (1.00 – 1.02) | (1.00 – 1.02) | (1.00 – 1.01) |
| IDACIn | 1.34 * | 1.34 * | 1.07 | 1.67 * | 1.26 | 1.29 |
| | (1.04 – 1.71) | (1.02 – 1.74) | (0.72 – 1.61) | (1.03 – 2.71) | (0.89 – 1.80) | (0.93 – 1.80) |
| white ethnicity percent | 1.00 *** | 0.99 *** | 1 | 0.99 | 1 | 1.00 * |
| | (0.99 – 1.00) | (0.99 – 1.00) | (1.00 – 1.00) | (0.99 – 1.00) | (0.99 – 1.00) | (0.99 – 1.00) |
| Year × IDACIn | 0.95 | 1.04 | 1.02 | 0.87 | 0.98 | 0.93 |
| | (0.83 – 1.09) | (0.88 – 1.23) | (0.79 – 1.30) | (0.66 – 1.16) | (0.79 – 1.22) | (0.76 – 1.13) |
| $Year^2$ × IDACIn | 1.01 | 0.99 | 0.99 | 1.02 | 1.01 | 1.01 |
| | (0.98 – 1.03) | (0.96 – 1.02) | (0.95 – 1.04) | (0.97 – 1.08) | (0.97 – 1.05) | (0.98 – 1.05) |
| Random Effects | | | | | | |
| $\sigma^2$ | 0.1 | 0.1 | 0.2 | 0.3 | 0.2 | 0.1 |
| $\tau_{00}$ | 0.1 $_{AreaName}$ | 0.1 $_{AreaName}$ | 0.3 $_{AreaName}$ | 0.4 $_{AreaName}$ | 0.2 $_{AreaName}$ | 0.2 $_{AreaName}$ |
| ICC | 0.6 | 0.6 | 0.6 | 0.6 | 0.6 | 0.6 |
| N | 143 $_{AreaName}$ | 143 $_{AreaName}$ | 143 $_{AreaName}$ | 143 $_{AreaName}$ | 143 $_{AreaName}$ | 143 $_{AreaName}$ |
| Observations | 659 | 662 | 662 | 662 | 661 | 662 |
| Marginal $R^2$ / Conditional $R^2$ | 0.133 / 0.688 | 0.186 / 0.646 | 0.009 / 0.579 | 0.049 / 0.623 | 0.102 / 0.623 | 0.106 / 0.630 |
| AIC | 438.395 | 629.381 | 1165.559 | 1371.251 | 989.879 | 900.755 |

* $p<0.05$ ** $p<0.01$ *** $p<0.001$

Table S14. Likelihood Ratio Tests of models including interaction effects of linear and quadratic Year x IDACI versus models without interaction effects

| Model | LogLik | Chi | p | df |
|---|---|---|---|---|
| I_asq | -180.19 | | | |
| Ii_asq | -179.43 | 1.5147 | 0.4689 | 2 |
| I_comm | -275.98 | | | |
| Ii_comm | -275.85 | 0.2579 | 0.879 | 2 |
| I_gm | -547.55 | | | |
| Ii_gm | -547.25 | 0.6084 | 0.7377 | 2 |
| I_fm | -652.13 | | | |
| Ii_fm | -651.43 | 1.4004 | 0.4965 | 2 |
| I_ps | -458.49 | | | |
| Ii_ps | -458.36 | 0.2522 | 0.8815 | 2 |
| I_pss | -413.55 | | | |
| Ii_pss | -413.26 | 0.5885 | 0.7451 | 2 |

Table S15. Number of LAs with outliers (Rate above 3 times the IQR from the upper IQR limit) by year

| Year | All five domains | Communication | Gross Motor | Fine Motor | Problem Solving | Personal social skills |
|---|---|---|---|---|---|---|
| 2018 | 2 | 4 | 6 | 5 | 5 | 5 |
| 2019 | 0 | 0 | 2 | 1 | 2 | 0 |
| 2020 | 1 | 2 | 3 | 1 | 3 | 1 |
| 2021 | 1 | 5 | 11 | 8 | 8 | 7 |
| 2022 | 3 | 4 | 4 | 6 | 5 | 5 |
| 2023 | 5 | 9 | 9 | 6 | 9 | 8 |
| 2024 | 2 | 5 | 1 | 2 | 3 | 3 |

Table S16. Models of the effect of IDACI controlled for ASQ coverage, ethnicity and year (no interaction effects), excluding outliers above 3 times the IQR

| | **All five domains** | **Communication** | **Gross Motor** | **Fine Motor** | **Problem Solving** | **Personal social skills** |
|---|---|---|---|---|---|---|
| *Predictors* | *Estimates* | *Estimates* | *Estimates* | *Estimates* | *Estimates* | *Estimates* |
| (Intercept) | 19.32 *** | 10.63 *** | 4.20 ** | 3.58 * | 2.99 ** | 5.86 *** |
| | (11.18 – 33.40) | (5.82 – 19.39) | (1.71 – 10.31) | (1.21 – 10.59) | (1.38 – 6.47) | (2.80 – 12.29) |
| Year [2020] | 1.05 | 1.07 | 0.94 | 0.96 | 1.07 | 1.01 |
| | (0.98 – 1.12) | (0.99 – 1.15) | (0.84 – 1.05) | (0.84 – 1.09) | (0.97 – 1.19) | (0.92 – 1.11) |
| Year [2021] | 1.08 * | 1.14 ** | 0.94 | 0.95 | 1.09 | 1.09 |
| | (1.01 – 1.16) | (1.05 – 1.24) | (0.83 – 1.06) | (0.82 – 1.10) | (0.98 – 1.21) | (0.99 – 1.21) |
| Year [2022] | 1.23 *** | 1.41 *** | 1.08 | 1.14 | 1.40 *** | 1.40 *** |
| | (1.15 – 1.32) | (1.30 – 1.53) | (0.96 – 1.21) | (1.00 – 1.31) | (1.26 – 1.55) | (1.27 – 1.54) |
| Year [2023] | 1.31 *** | 1.41 *** | 1.15 * | 1.27 *** | 1.55 *** | 1.50 *** |
| | (1.23 – 1.41) | (1.30 – 1.53) | (1.02 – 1.29) | (1.11 – 1.46) | (1.40 – 1.72) | (1.36 – 1.66) |
| Year [2024] | 1.28 *** | 1.35 *** | 1.13 * | 1.21 ** | 1.51 *** | 1.42 *** |
| | (1.19 – 1.37) | (1.24 – 1.45) | (1.01 – 1.26) | (1.06 – 1.39) | (1.37 – 1.67) | (1.29 – 1.56) |
| IDACIn | 1 | 1 | 1 | 1 | 1 | 1 |
| | (0.99 – 1.00) | (0.99 – 1.01) | (0.99 – 1.01) | (0.99 – 1.01) | (1.00 – 1.01) | (0.99 – 1.01) |
| COV Value | 1.23 * | 1.39 ** | 1.06 | 1.36 | 1.33 * | 1.21 |
| | (1.01 – 1.50) | (1.14 – 1.69) | (0.79 – 1.42) | (0.93 – 1.98) | (1.03 – 1.71) | (0.95 – 1.54) |
| white ethnicity percent | 1.00 ** | 0.99 *** | 1 | 1 | 1 | 1 |
| | (0.99 – 1.00) | (0.99 – 1.00) | (1.00 – 1.01) | (0.99 – 1.00) | (0.99 – 1.00) | (0.99 – 1.00) |
| **Random Effects** | | | | | | |
| $\sigma^2$ | 0.1 | 0.1 | 0.2 | 0.2 | 0.1 | 0.1 |
| $\tau_{00}$ | 0.1 AreaName | 0.1 AreaName | 0.2 AreaName | 0.4 AreaName | 0.2 AreaName | 0.1 AreaName |
| ICC | 0.6 | 0.5 | 0.6 | 0.6 | 0.5 | 0.5 |
| N | 143 AreaName | 143 AreaName | 143 AreaName | 143 AreaName | 143 AreaName | 143 AreaName |
| Observations | 652 | 648 | 643 | 646 | 641 | 645 |
| Marginal $R^2$ / Conditional $R^2$ | 0.134 / 0.684 | 0.197 / 0.632 | 0.019 / 0.561 | 0.054 / 0.619 | 0.127 / 0.593 | 0.118 / 0.598 |
| AIC | 409.361 | 552.986 | 1039.016 | 1278.021 | 862.517 | 813.251 |

* *p<0.05* ** *p<0.01* *** *p<0.001*

**Appendix S5. Supplemental Figures**

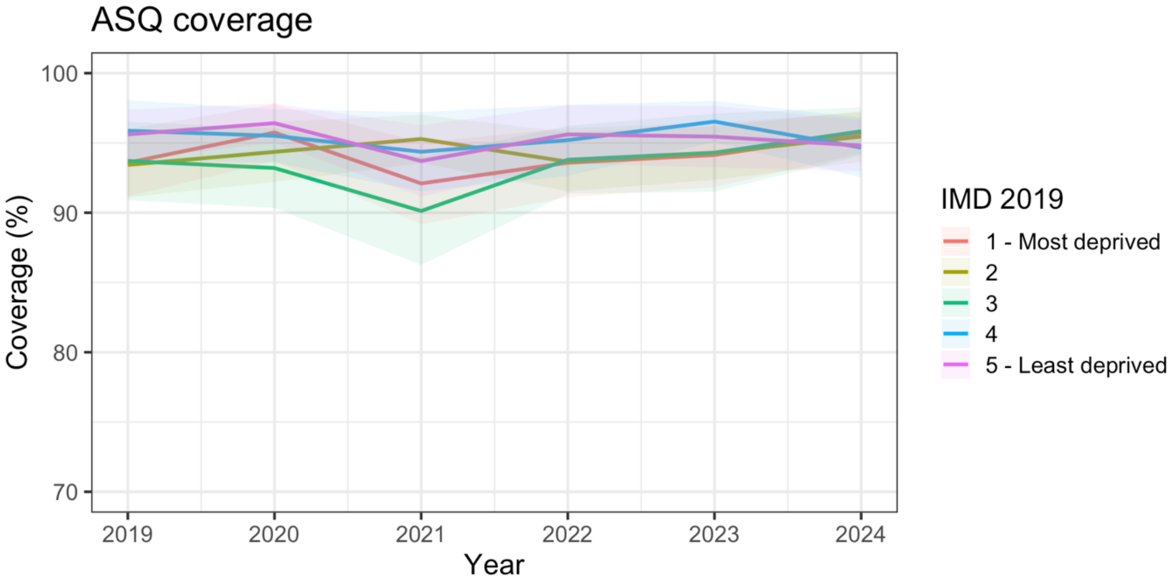


Figure S1. Time trends in the ASQ coverage by IMD quintile

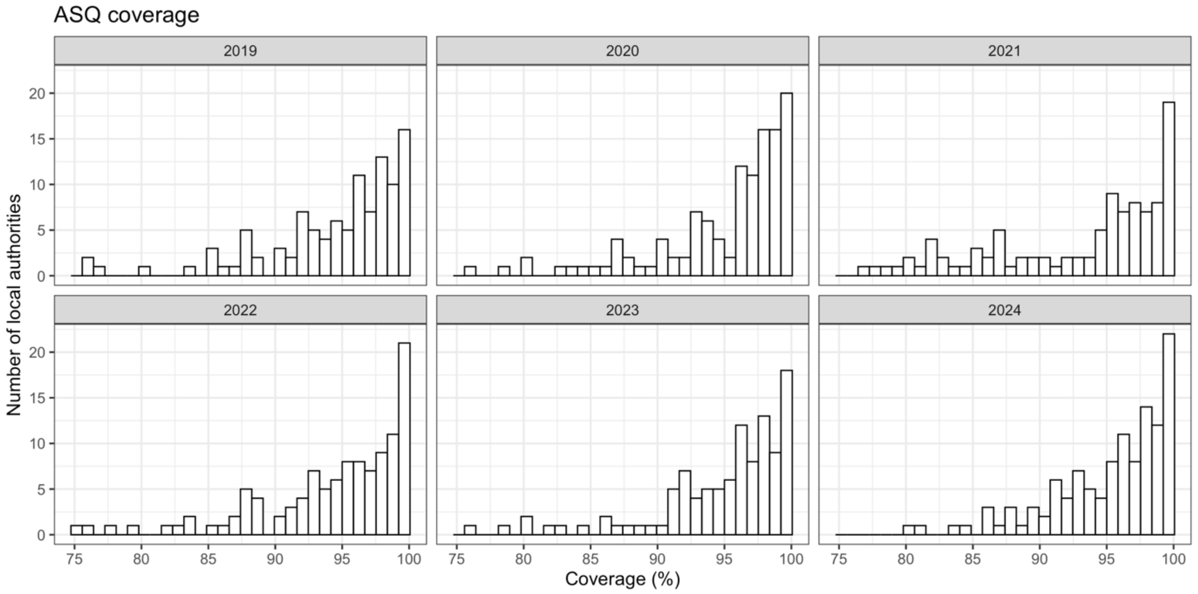


Figure S2. Histogram showing variation in coverage for the analysis sample (>75% coverage)

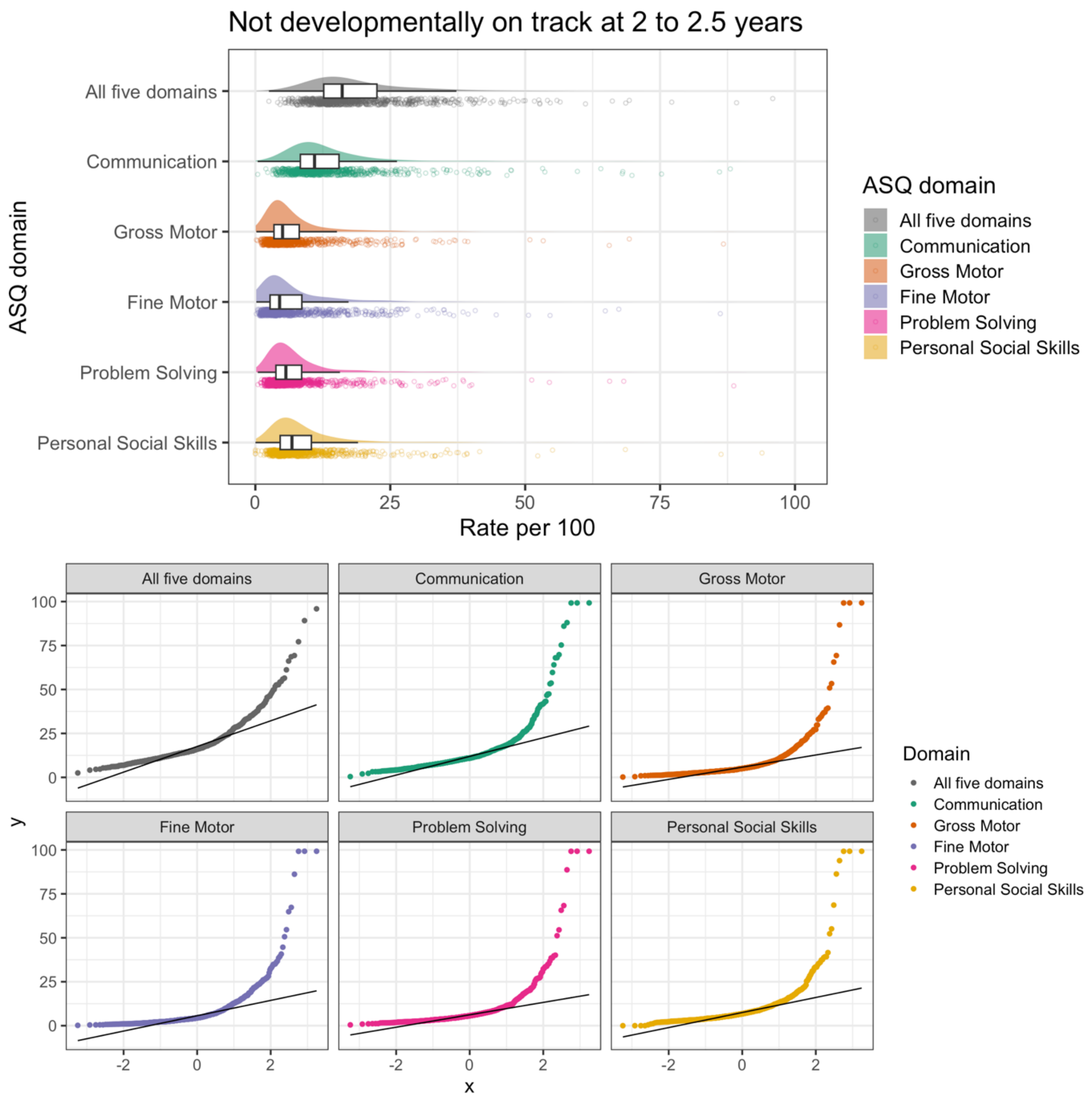


Figure S3. Raincloud plot (top) and normal q-q plot (bottom) showing the positive skew of the outcome in the original scale

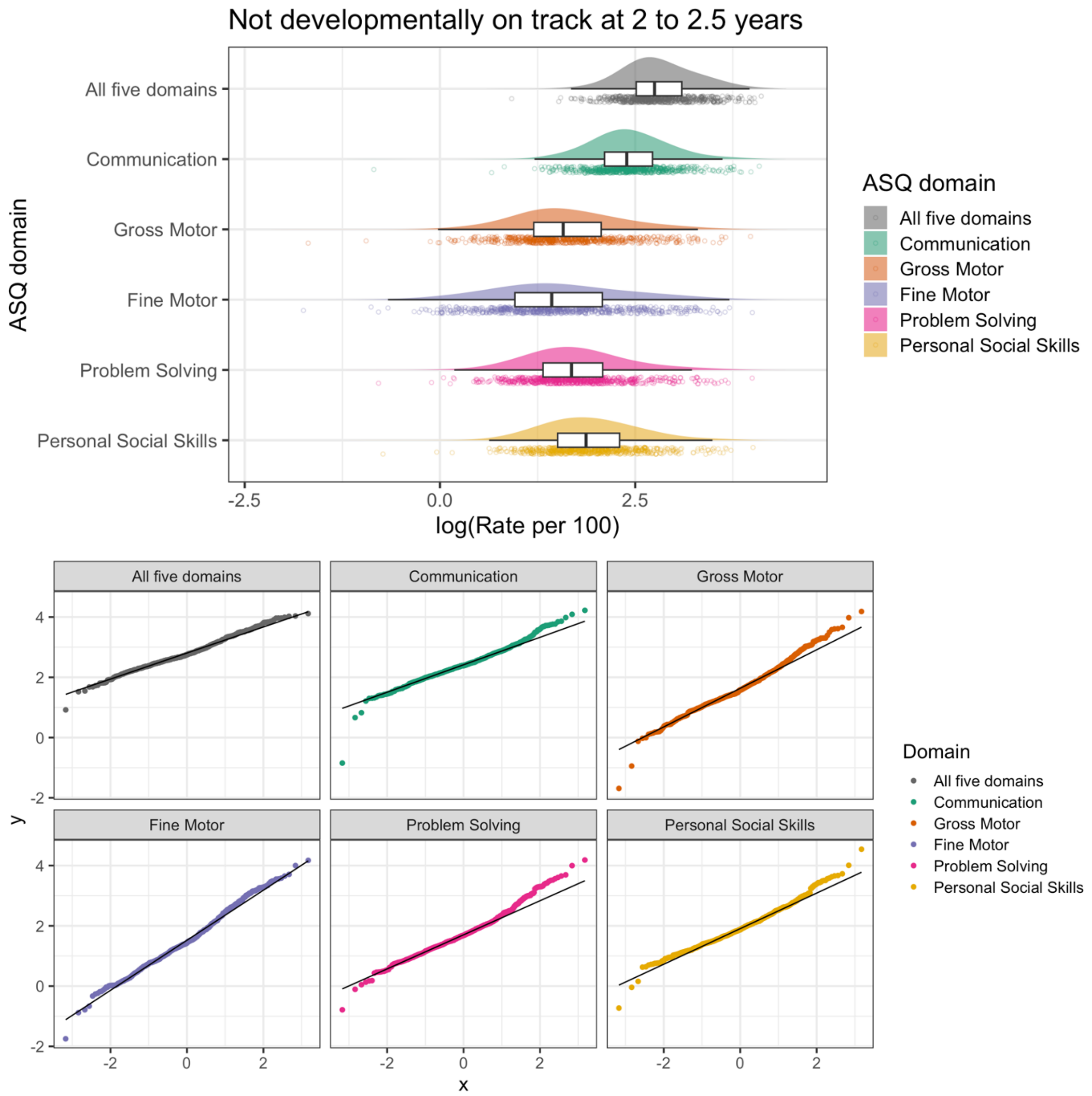


Figure S4. Raincloud plot (top) and normal q-q plot (bottom) of the log-transformed outcome showing it approximates a normal distribution

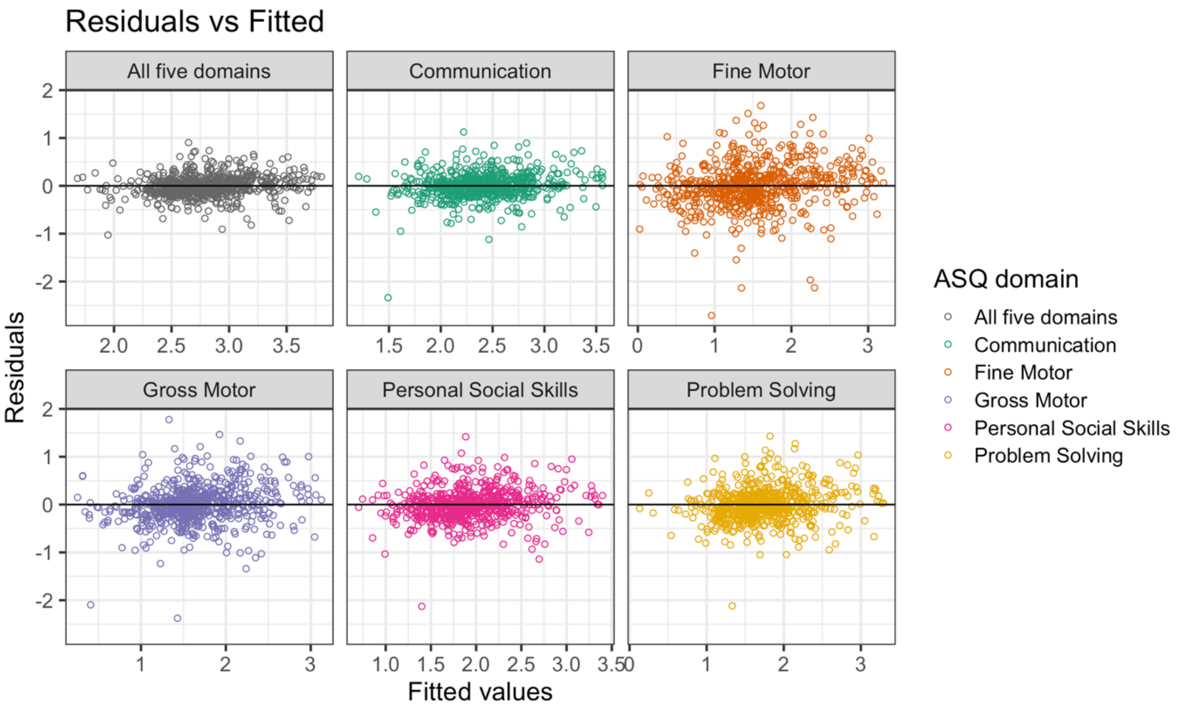


Figure S5. Residual diagnostics for models using the log-transformed outcome. No trend in the residuals against fitted values or the variance in the residuals across different fitted values

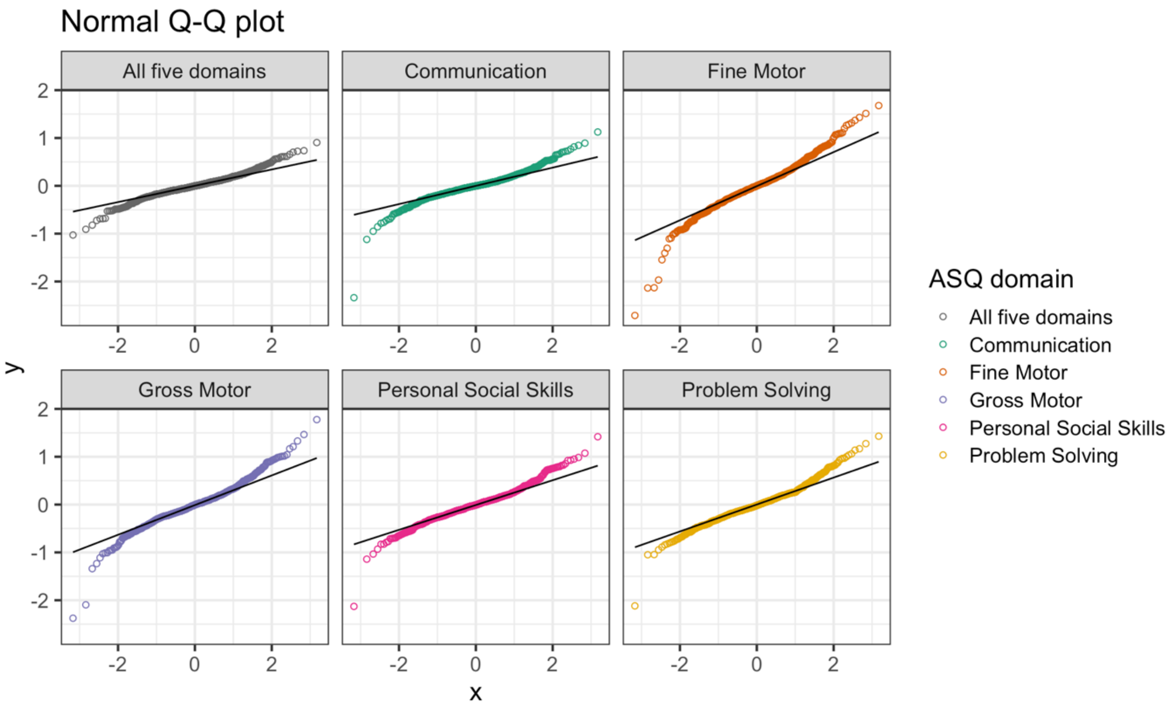


Figure S6. Residual diagnostics for models using the log-transformed outcome. Residuals are normally distributed

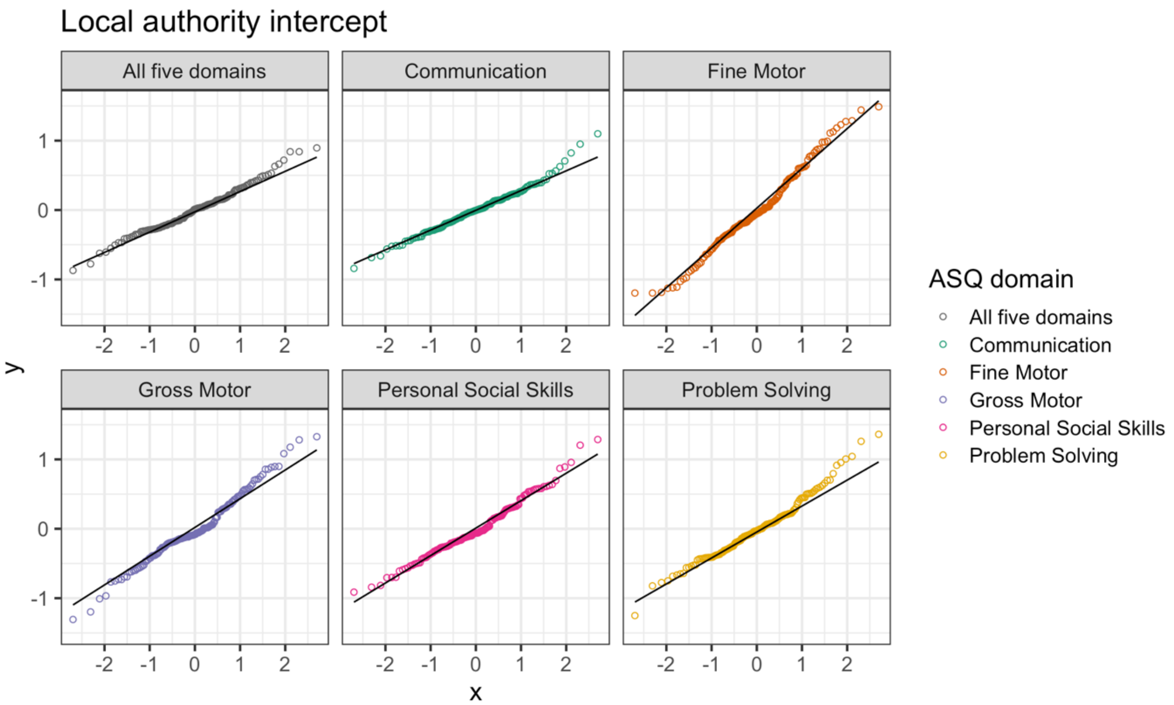


Figure S7. Model diagnostics. Random effects are correctly specified (intercepts for each local authority follows a normal distribution

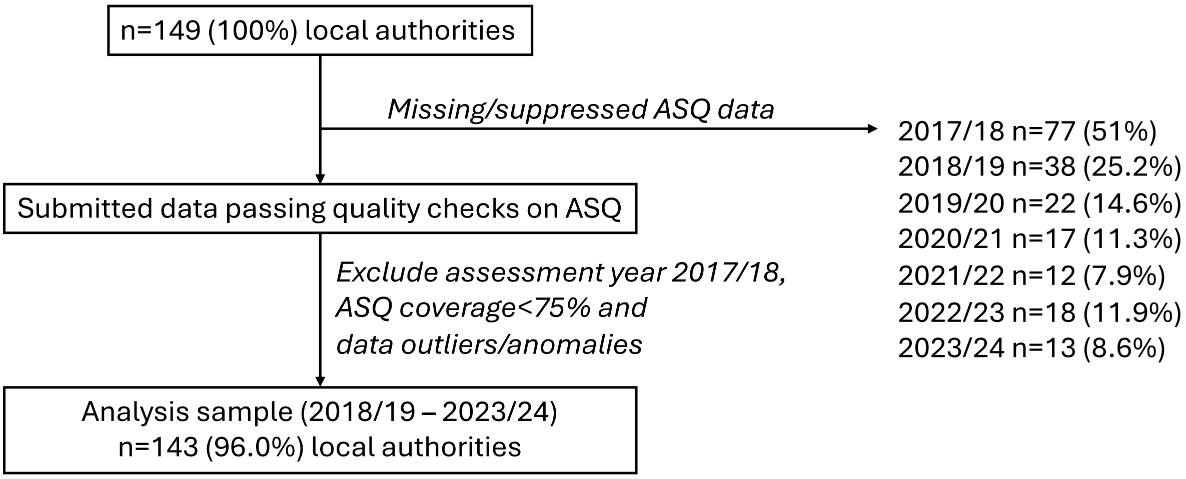


Figure S8. Inclusion and exclusion criteria for analysis

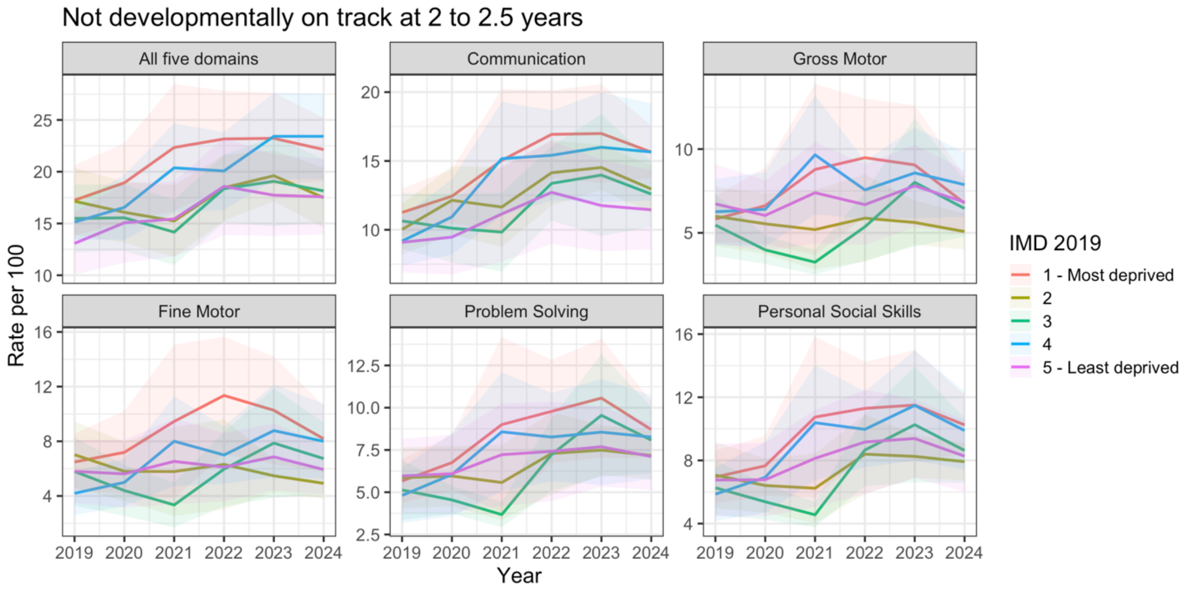


Figure S9. Time trends in rates of developmental outcome by IMD quintile (means and 95% confidence intervals provided)
